# Targeted Long-Read sequencing provides functional validation of variants predicted to alter splicing

**DOI:** 10.64898/2026.03.02.26346984

**Authors:** Ilaria Quartesan, Arianna Manini, Ricardo Parolin Schnekenberg, Stefano Facchini, Riccardo Curro, Arianna Ghia, Alessandro Bertini, James Polke, Enrico Bugiardini, Pinki Munot, Mary O’Driscoll, Matilde Laurá, James N. Sleigh, Mary M Reilly, Henry Houlden, Nicholas Wood, Andrea Cortese

## Abstract

**Background:** Whole-genome sequencing (WGS) has improved the diagnosis of rare genetic disorders, yet interpretation of non-coding variants that affect splicing remains challenging. *In silico* predictions alone are insufficient, and short-read RNA sequencing may fail to capture complex or low-abundance splicing events. Targeted amplicon-based long-read RNA sequencing (Amp-LRS) offers a cost-effective approach for functional validation of candidate splice-altering variants.

**Methods:** We applied Amp-LRS to five patients with neurological disorders (central nervous system, peripheral nervous system, or muscle) harbouring candidate non-coding variants predicted to alter splicing. RNA was extracted from fibroblasts or peripheral blood, and full-length transcript amplicons were sequenced using Oxford Nanopore Technologies. Nonsense-mediated decay (NMD) inhibition was performed on fibroblast cultures using cycloheximide.

**Results:** Amp-LRS validated all five candidate variants, including intronic and UTR variants in *POLR3A, OPA1, PYROXD1, GDAP1*, and *SPG11*. Aberrant splicing events included exon skipping, intron retention, cryptic splice site activation, and pseudoexon inclusion, often resulting in frameshifts and premature termination codons. For *POLR3A* and *OPA1*, multiple abnormal isoforms arose from single variants, highlighting the complexity of splicing disruption. Some pathogenic effects were detectable only in a minority of reads and variably enriched by NMD inhibition, consistent with being hypomorphic. The approach was successfully applied using accessible tissues and enabled multiplexed sequencing at low per-sample cost.

**Conclusions:** Amp-LRS is a sensitive, versatile, and cost-effective method for functional assessment of non-coding splice-altering variants identified by WGS. By enabling full-length transcript analysis from accessible tissues, this approach improves interpretation of variants of uncertain significance and could enhance molecular diagnosis in rare neurological diseases.

## 1. Introduction

Whole-Genome Sequencing (WGS) has transformed the diagnosis of genetic disorders, yet interpreting variants of unknown significance (VUS), especially those outside coding regions, remains a major challenge in clinical genomics. More specifically, variants in introns and untranslated regions (UTRs) that disrupt splicing are increasingly recognised as important contributors to Mendelian diseases, but *in silico* tools cannot reliably predict their functional consequences [1]. RNA sequencing has therefore emerged as a powerful complementary diagnostic tool, with studies demonstrating increased diagnostic yields of 10-35% when applied to previously undiagnosed cohorts [2], [3].

More recently, long-read sequencing (LRS) has been applied to transcriptome analysis, enabling full-length transcript capture and allele phasing not achievable with short-read approaches [4], [5]. Nevertheless, the cost of whole-transcriptome LRS currently limits its routine clinical implementation. Targeted amplicon-based long-read RNA sequencing (Amp-LRS) represents an affordable method to analyse splicing effects within full-length transcripts while retaining allele phasing information.

In this study, we applied Amp-LRS to five patients with neurological disorders in whom short-read WGS had identified candidate splice-altering variants in genes consistent with their clinical presentations.

## 2. Methods

### 2.1 Short-read Whole-Genome Sequencing

Short-read WGS was performed on DNA extracted from peripheral blood.as part of the UK 100,000 Genomes Project, a national research initiative led by Genomics England in partnership with the National Health Service. Variants were filtered and prioritised based on predicted functional impact, population frequency, and consistency with the clinical phenotype. *SpliceAI* was used to predict the effect of candidate variants on splicing [6].

### 2.2 Amplicon-based Full-length Transcript Sequencing

Fibroblasts were cultured in Dulbecco’s Modified Eagle Medium (DMEM) with 10% Fetal Bovine Serum (FBS). RNA was extracted from ∼80% confluent fibroblast cultures derived from Patients 1-3 using the RNeasy Kit (Qiagen) according to the manufacturer’s instructions. For Patients 4 and 5, RNA was extracted from whole blood stabilised in Tempus Blood RNA tubes using the Tempus Spin RNA Isolation Kit (Applied Biosystems). cDNA was synthesised using random hexamers and SuperScript III Reverse Transcriptase (Invitrogen). Full-length transcripts of each gene of interest were PCR-amplified from cDNA in 50 µL PCR reactions of Phusion High-Fidelity PCR Master Mix (Thermo Scientific) using primers annealing to the 5’ and 3’ UTRs of the MANE Select transcript of each gene (***Table 1***). For *SPG11*, full-length transcript amplification was not feasible due to the large cDNA size (∼9.5 kb). A targeted amplicon strategy was therefore employed using primers flanking the region of interest (*i*.*e*., exons 11–13).

**Table 1:**
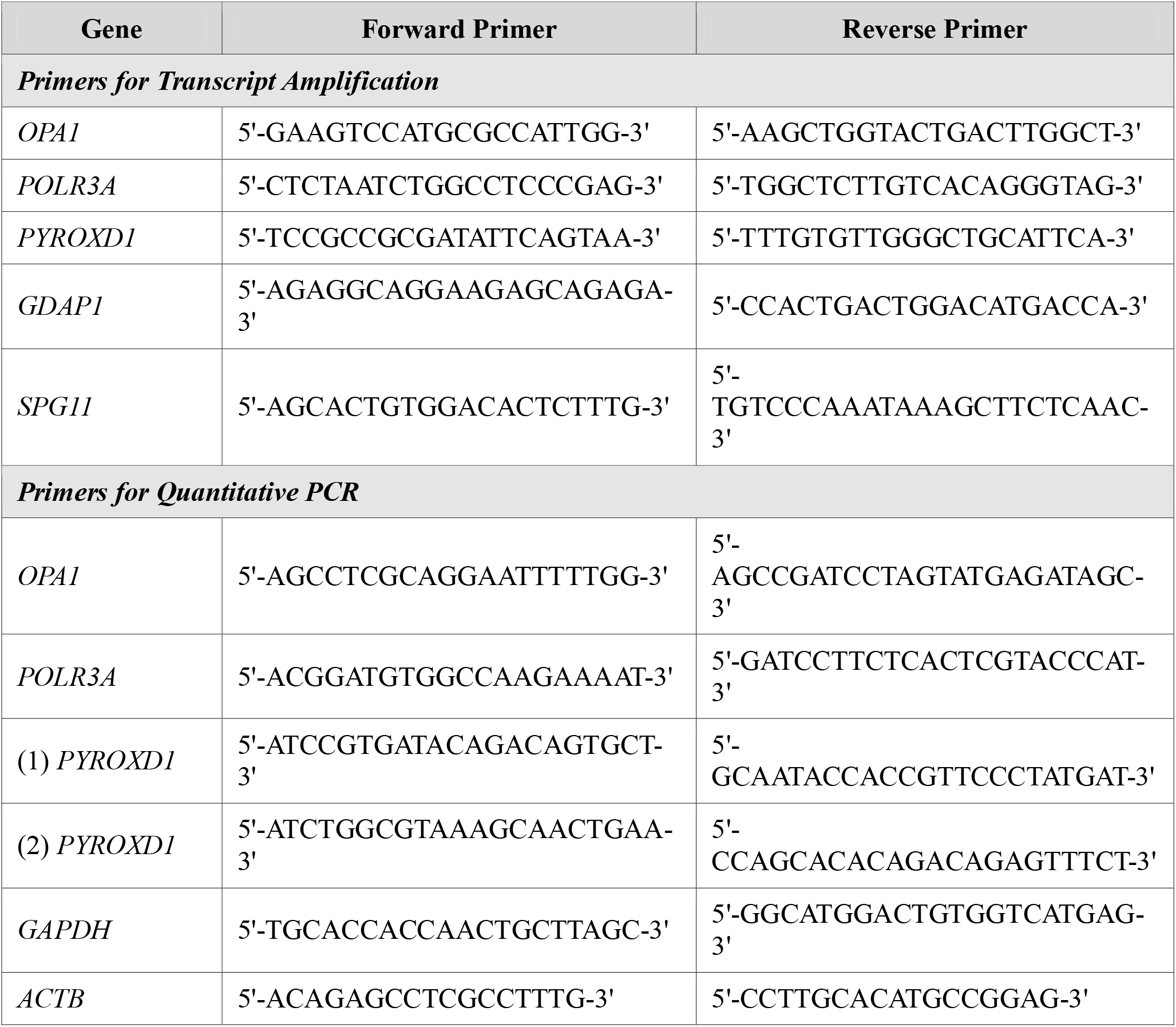
Oligonucleotide primer sequences used for full-length transcript amplification and quantitative real-time PCR (RT-qPCR) analysis.

For Patients 1-3 and their controls, amplicons were prepared for long-read sequencing with the Ligation Sequencing DNA V14 kit (SQS-LSK114, Oxford Nanopore Technologies [ONT]) and sequenced on a Flongle flow cell (ONT) over 24 h. Amplicons from three controls were pooled in equimolar ratios and sequenced in the same flow cell, without barcoding. For Patients 4 and 5 and their matched control, amplicons were bead-purified and sequenced as PCR libraries by a commercial provider (Full Circle Labs, London, UK).

Sequencing reads were aligned to the GRCh38 reference genome using minimap2 [7]. Alignments were visually inspected using Integrative Genomic Viewer (IGV) [8]. To quantify transcript isoforms, read depth was calculated from aligned BAM files using mosdepth, restricting analysis to a minimum mapping quality of 30 [9].

### 2.3 Nonsense-Mediated Decay (NMD) Inhibition Assay

To assess the effect of NMD on aberrant transcripts, cultured fibroblasts from Patients 1–3 were treated with cycloheximide prior to RNA extraction. At 90–100% confluency, cells were dissociated with 1.5 mL TrypLE Express (Gibco) for 8 minutes at 37°C, counted on a Countess II automated cell counter (Invitrogen), and seeded into 6-well plates at 1.25 × 10□ or 2.5 × 10□ cells per well. After 24 hours, culture medium was replaced with DMEM/5% FBS containing either 100 µg/mL cycloheximide (Sigma-Aldrich) or 0.1% DMSO for 6 hours at 37°C. Cells were then harvested, pelleted by centrifugation at 1000 × *g* for 5 minutes, snap-frozen on dry ice, and stored at −80°C until RNA extraction.

### 2.4 Quantification of Gene Expression

For Patients 1-3, quantitative PCR (qPCR) was performed using FAST SYBR Green Master Mix (Applied Biosystems) with custom primers (***Table 1***) and measured with a QuantStudio 7 Flex Real-Time PCR System (Applied Biosystems). Relative gene expression was calculated using the comparative Ct (2^−ΔΔCt) method. Target gene expression was normalized to the geometric mean of two reference genes, ACTB and GAPDH. Relative expression was calculated using qRAT software and presented as fold expression change [10]. Comparative analyses for each gene were conducted between each patient and three controls.

## 3. Results

### 3.1 Clinical Phenotypes and Variant Identification

Five patients were assessed a tertiary neurogenetics referral centre, and investigated with short-read WGS. After filtering and prioritisation, variants in genes consistent with the phenotypes of patients were selected for further investigations. Detailed clinical features and genetic findings are presented in ***Table 2***.

**Table 2:**
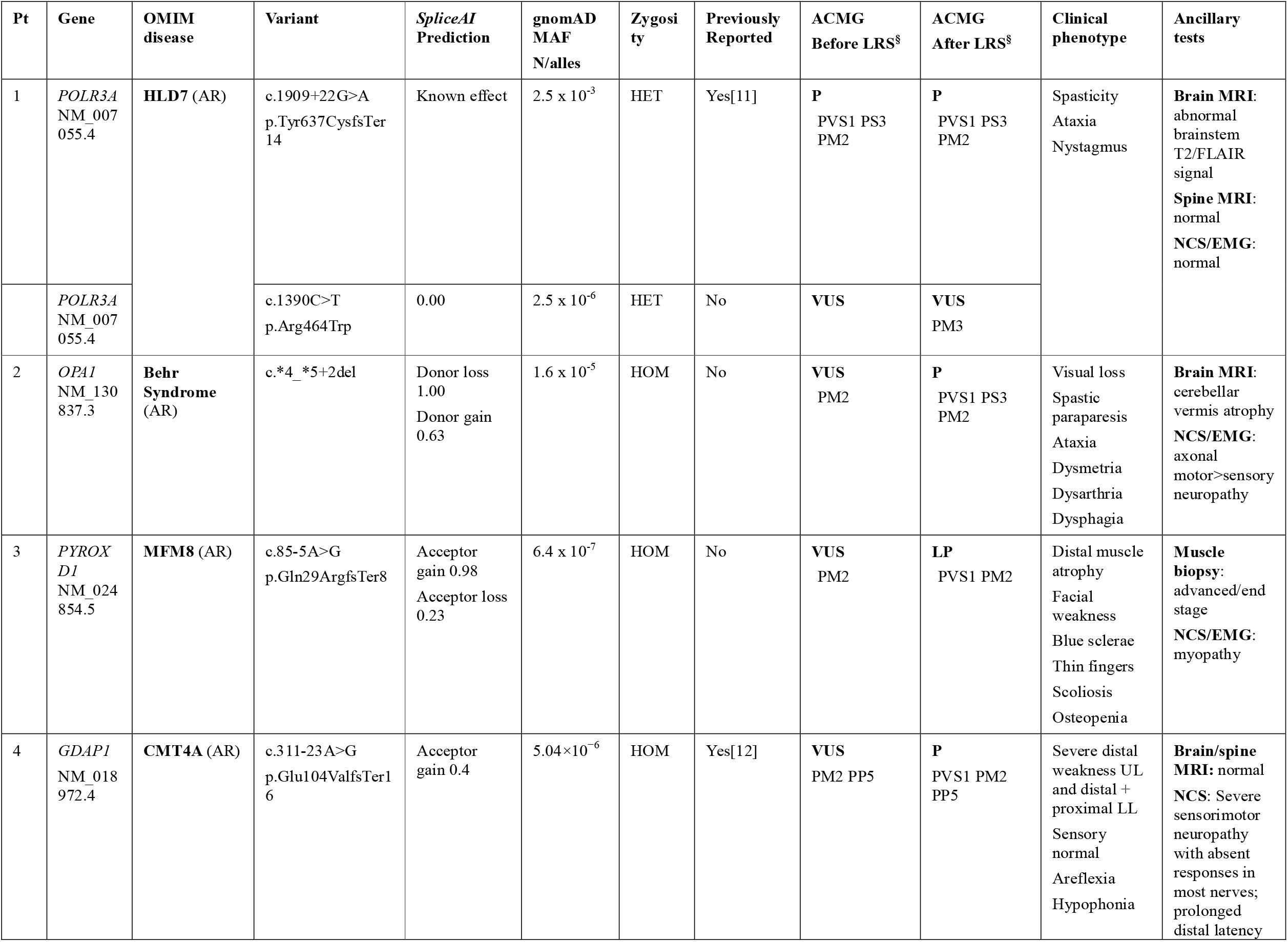
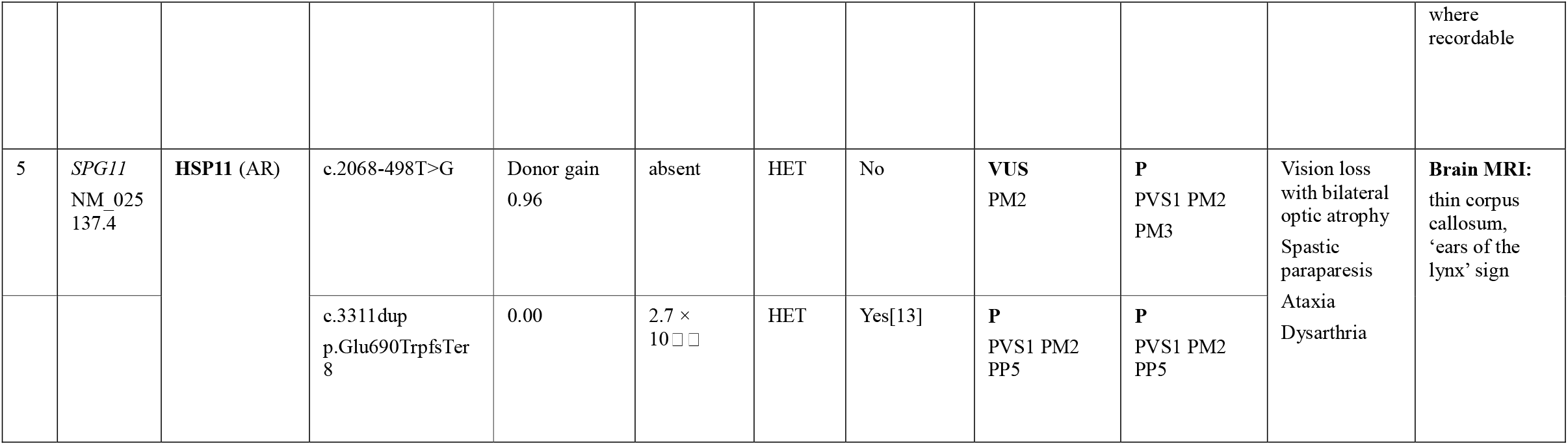
Clinical and genetic features of patients with candidate splicing variants investigated by targeted long-read RNA sequencing. ^†^Reference genome GRCh38. ^§^The variant classification according to the American College of Medical Genetics and Genomics (ACMG) guidelines was assessed before and after performing targeted long-read sequencing of the full-length transcripts. AR: autosomal recessive; EMG: electromyography; F: female; fs: frameshift; HET: heterozygous; HLD7: Leukodystrophy, hypomyelinating, 7, with or without oligodontia and/or hypogonadotropic hypogonadism; HOM: homozygous; LL: lower limbs; LRS: long-read sequencing; M: male; MAF: minor allele frequency; MFM8: Myofibrillar myopathy type 8; MRI: magnetic resonance imaging; NA: not available; NCS: nerve conduction studies; Pt: patient; UL: upper limbs;↓: reduced; ↑: increased.

Patient 1 was investigated for adolescent-onset progressive spastic ataxia and was found to harbour two heterozygous variants in *POLR3A*: a previously reported pathogenic intronic mutation (c.1909+22G>A) [11] and a novel missense variant (c.1390C>T; p.Arg464Trp) ***(Figure 1A)***.

**Figure 1.**
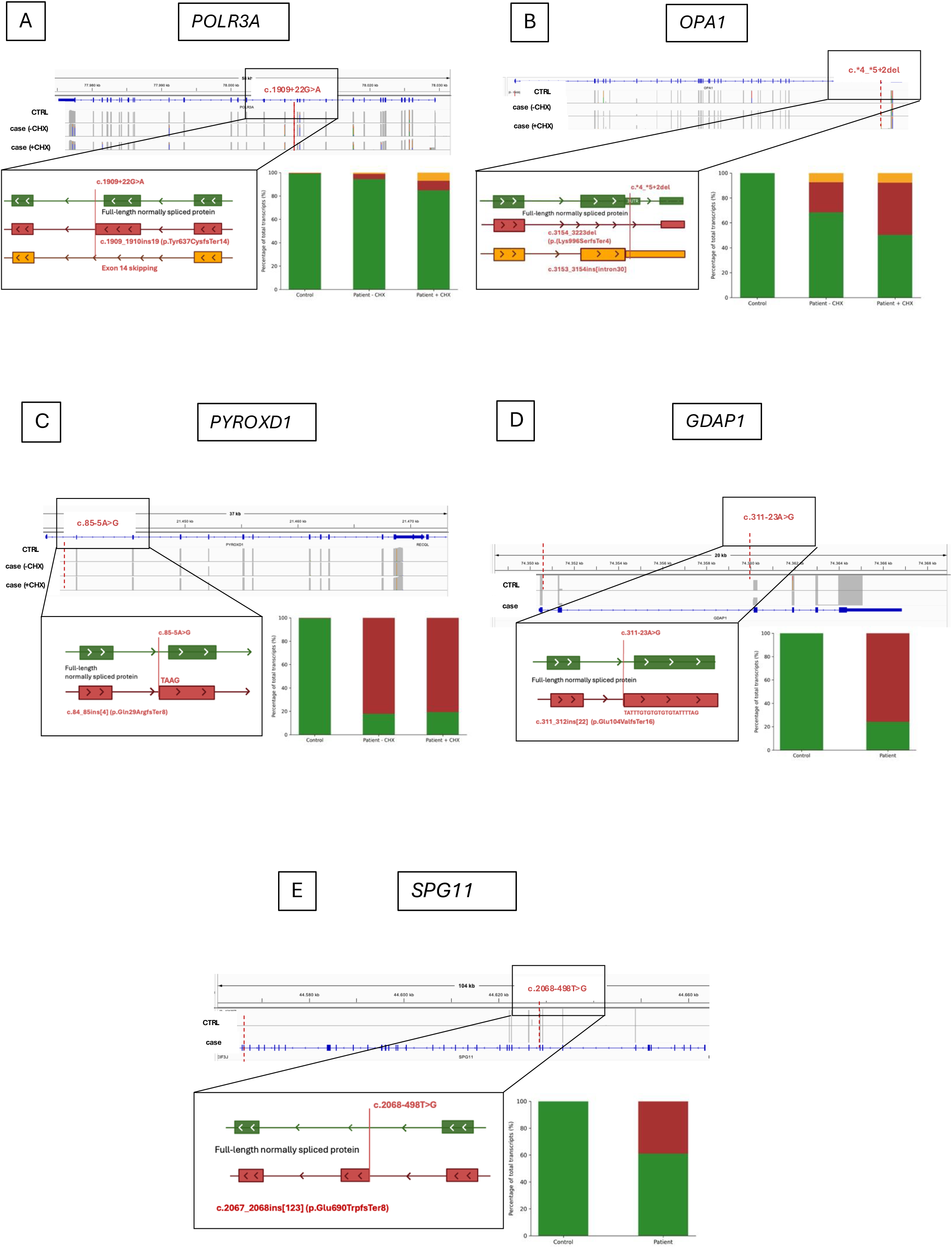
Identification and functional characterisation of splice-altering variants using targeted long-read RNA sequencing (Amp-lrRNAseq). Five patients with neurogenetic disorders harbouring candidate splicing variants were investigated using targeted Amp-LRS. Each panel (A–E) shows: Integrative Genomics Viewer (IGV) visualisation of long-read alignments at the splice junction of interest, comparing control sample (CTRL), patient sample without cycloheximide treatment (case −CHX), and patient sample after NMD inhibition with cycloheximide (case +CHX) where applicable; a schematic representation of the splicing defect, depicting the wild-type transcript (top) and the aberrant transcript(s) caused by the variant (bottom), with the predicted protein consequence; and a bar chart showing the quantification of aberrant transcripts as a percentage of total reads. **(A)** *POLR3A* c.1909+22G>A: this deep intronic variant activates a cryptic splice donor site, resulting in retention of the first 19 nucleotides of intron 14 and leading to a frameshift (p.Tyr637CysfsTer14). Additional exon 14 skipping was also observed. **(B)** *OPA1* c.*4_*5+2del: this 4-bp deletion disrupts the splice donor site of the penultimate exon located in the 3′ UTR. **(C)** *PYROXD1* c.85-5A>G: this intronic variant creates a cryptic splice acceptor site, resulting in the inclusion of four intronic nucleotides (ATAG) at the 5′ end of exon 2, causing a frameshift (p.Gln29ArgfsTer8). **(D)** *GDAP1* c.311-23A>G: this deep intronic variant activates a cryptic splice acceptor site within intron 2, leading to the inclusion of 22 intronic nucleotides and a frameshift (p.Glu104ValfsTer16). **(E)** *SPG11* c.2068-498T>G: this deep intronic variant creates a novel splice donor site, resulting in the insertion of a 105-bp pseudoexon between exons 11 and 12, leading to a frameshift (p.Glu690TrpfsTer8).

Patient 2 presented with early-onset optic atrophy, cerebellar ataxia, pyramidal signs and neuropathy consistent with Behr syndrome. Genetic analysis revealed a novel homozygous 4-bp deletion affecting the 3’ UTR splice donor site of *OPA1* (c.*4_*5+2del) ***(Figure 1B)***.

Patient 3, investigated for skeletal abnormalities and early-onset myopathy, harboured a novel homozygous intronic variant in *PYROXD1* (c.85-5A>G) ***(Figure 1C)***.

Patient 4 presented with childhood-onset severe predominantly motor axonal neuropathy and hypophonia secondary to vocal cord paresis. A homozygous deep intronic variant in *GDAP1* (c.311-23A>G), previously reported in an Iranian family with axonal CMT [12], was identified ***(Figure 1D)***.

Patient 5 and an affected sibling presented in early adulthood with spastic paraparesis, ataxia, dysarthria, and visual impairment. MRI showed thin corpus callosum and ear of the lynx sign. Compound heterozygous variants in *SPG11* were identified: a known pathogenic frameshift mutation (c.3311dup) [13] and a novel deep intronic variant of uncertain significance (c.2068-498T>G) ***(Figure 1E)***.

No additional pathogenic or likely pathogenic variants were detected in any patient that could explain their clinical presentations.

### 3.2 Validation of Amp-LRS with known POLR3A variant

The heterozygous *POLR3A* c.1909+22G>A variant, previously reported in compound heterozygosity with another *POLR3A* mutation, is associated with ataxia and spasticity, closely matching the clinical features of Patient 1 [11]. Previous studies using PCR and Sanger sequencing showed that this hypomorphic variant activates a ‘leaky’ cryptic splice site in intron 14 [11].

Quantitative analysis of read depth, calculated using mosdepth, showed that the first 19 nucleotides of intron 14 were retained as part of the mature transcript. In the untreated patient sample, this aberrant retention accounted for ∼1.1% of the total coverage at this locus, compared to 0.4% in controls ***(Figure 1A)***. Partial intron 14 retention results in a frameshifted transcript that, if translated, is expected to cause premature protein truncation (p.Tyr637CysfsTer14). Following cycloheximide treatment to inhibit NMD, the proportion of aberrant transcripts increased to 7.0%, representing a 6.4-fold enrichment compared to the untreated state.

Notably, Amp-LRS revealed a more complex disruption of splicing associated with this variant, including skipping of exon 14 in 4.4% of patient reads (increasing to 8.1% after cycloheximide treatment), compared to ∼0.4% in controls.

qPCR showed a small (∼13%), non-significant reduction in *POLR3A* expression in the patient fibroblasts relative to controls.

### 3.3 Investigation of novel variants with Amp-lrRNAseq

Patients 2-5 carried variants predicted by *SpliceAI* to alter pre-mRNA splicing (***Table 1***). To assess their functional impact, Amp-LRS was performed on RNA derived from patient-specific fibroblast cultures (Patients 2 and 3) or peripheral blood (Patients 4 and 5). NMD inhibition was applied when fibroblasts were available to facilitate detection of unstable aberrant transcripts.

#### 3.3.1 OPA1

The homozygous variant identified in Patient 2 is a novel 4-nucleotide deletion affecting the splice donor site of the penultimate exon of *OPA1*, located downstream of the canonical stop codon in the 3’ UTR (NM_130837.3:c.*4_*5+2del) ***(Figure 1B)***.

Amp-LRS revealed that this variant significantly impacts splicing at the 3’ end of *OPA1* transcripts. Quantitative analysis of read depth showed that ∼24.1% of reads from untreated patient cells showed skipping of the last coding exon (exon 30). Following NMD inhibition, the proportion of exon 30-skipping transcripts increased to 41.8%, indicating that these aberrantly spliced transcripts are subject to NMD. Consistent with this, qPCR analysis demonstrated a ∼20% reduction in *OPA1* expression in patient fibroblasts relative to controls (p=0.039) (*Figure 2B*).

In addition to exon skipping, isoform-level analysis identified a smaller subset (7.4%) of *OPA1* transcripts characterised by retention of intron 30 downstream of the last exon containing the canonical stop codon. These intron-retaining isoforms showed minimal change following NMD inhibition (7.8%), suggesting they may escape NMD surveillance due to the absence of a downstream exon-exon junction [14].

#### 3.3.2 PYROXD1

The homozygous *PYROXD1* (NM_024854.5) c.85-5A>G variant in Patient 3 was predicted by *SpliceAI* to create a cryptic acceptor site (delta score 0.98) proximal to the canonical intron 1-exon 2 boundary ***(Figure 1C)***.

Amp-LRS confirmed this prediction, demonstrating that the variant introduces an alternative splice acceptor site that results in inclusion of four intronic nucleotides (ATAG) at the 5’ end of exon 2. This aberrant splicing event was identified in 82.1% of the reads from patient fibroblasts, compared to 0.45% in control samples. The insertion causes a frameshift leading to a p.Gln29ArgfsTer8 premature termination codon (PTC) that, if translated, would result in a truncated protein lacking both enzymatic domains of the *PYROXD1* oxidoreductase [15].

Following NMD inhibition, the proportion of reads supporting the aberrant transcript remained unchanged (80.6%). Moreover, *PYROXD1* expression was similar between cycloheximide-treated and untreated cells. These results suggest that, despite the expected functional severity of the c.85-5A>G variant, aberrantly spliced *PYROXD1* transcripts may partly escape NMD and generate a non-functional truncated protein. This is likely explained by PTC proximity to the start codon, as PTCs within the first ∼200 nucleotides frequently escape NMD surveillance [16], a phenomenon documented for other first-exon variants including the *BRCA1*185delAG Ashkenazi founder mutation [17].

#### 3.3.3 GDAP1

The homozygous *GDAP1* (NM_018972.4) c.311-23A>G variant in Patient 4 had previously been reported in Iranian families with axonal CMT, where standard RT-PCR and Sanger sequencing from blood suggested partial intron retention [12].

Our Amp-LRS analysis accurately characterised the splicing defect, confirming that the variant activates a cryptic splice acceptor site within intron 2 ***(Figure 1D)***. Quantitative analysis of read depth showed that this aberrant splicing event resulted in the inclusion of the last 22 nucleotides of intron 2, which was detected in 75.9% of patient reads. In contrast, this aberrant transcript was absent in the control sample (<0.02%). The 22-nucleotide insertion causes a frameshift and introduces a premature stop codon (p.Glu104ValfsTer16), predicted to trigger NMD and result in loss of function.

#### 3.3.4 SPG11

The novel *SPG11* (NM_025137.4): c.2068-498T>G variant was identified in Patient 5 in compound heterozygosity with the known pathogenic frameshift mutation c.3311dup. Located 498 nucleotides upstream of exon 12, this deep intronic variant was predicted by *SpliceAI* to create a novel splice donor site (delta score 0.96). Amp-LRS analysis from patient blood identified an abnormal splicing event caused by this variant: the insertion of a 105-bp pseudoexon derived from intronic sequence between exons 11 and 12 of the mature transcript ***(Figure 1E)***. This pseudoexon insertion is predicted to lead to a frameshift by introducing a premature stop codon (p.Glu690TrpfsTer8). 38.9% of *SPG11* transcripts in the patient sample harboured the abnormal pseudoexon, while no such event was detected in control samples.

## 4. Discussion

Interpretation of non-coding genetic variants is a growing challenge in the era of WGS. Our study demonstrates that Amp-LRS of accessible patient-derived blood and skin fibroblast tissues is an effective method for the functional assessment of non-coding variants predicted to affect splicing.

Using this targeted long-read RNA sequencing approach, we provided functional validation of splice-altering variants across a spectrum of neurological disorders affecting the central nervous system, peripheral nervous system, and muscle. Compared with whole-transcriptome long-read RNA sequencing, the amplicon-based approach substantially reduces costs. Once gene-specific amplicons are generated, long-read sequencing can be multiplexed using barcoded primers, with a per-sample sequencing cost of ∼£10 (∼$12 USD), making this approach compatible with routine diagnostic workflows.

Beyond confirming predicted splice defects, Amp-LRS enabled analysis of full-length transcripts for most genes, revealing complex and sometimes unexpected splicing consequences. In *POLR3A* and *OPA1*, we identified multiple aberrant isoforms arising from single non-coding variants, including exon skipping and intron retention events, underscoring that the impact of splice-altering variants is often broader than a single, isolated splicing outcome. This complexity is often overlooked by conventional short-read RNA sequencing or targeted RT-PCR approaches, which typically focus on predefined exon–exon junctions [18].

A key observation across several cases was that disease-relevant aberrant transcripts were sometimes present in only a small fraction of reads and were only partially enriched following NMD inhibition. This is not unexpected, as many splice-altering variants exert hypomorphic effects rather than complete loss of function. The *POLR3A* c.1909+22G>A variant illustrates this principle well: disease severity in *POLR3A*-related disorders correlates with residual protein function, ranging from severe neonatal Wiedemann-Rautenstrauch syndrome (biallelic truncating variants) [19], through early-onset 4H leukodystrophy [20], to later-onset spastic ataxia caused by the deep intronic c.1909+22G>A variant [11]. These findings emphasise that low-abundance aberrant transcripts can nevertheless be clinically meaningful and should not be dismissed in functional analyses.

Our findings also provide insight into the performance of *in silico* splicing prediction tools. *SpliceAI* correctly prioritised all variants of uncertain significance investigated in this study and accurately predicted the primary splicing defect in most cases, including the *GDAP1* c.311-23A>G variant, which despite a relatively modest delta score (0.4) produced aberrant transcripts in over 75% of reads. However, false-negative predictions remain an important limitation, as illustrated by the *POLR3A* c.1909+22G>A variant, which is known to be pathogenic from previous studies and from our own functional data, but received a relatively low *SpliceAI* score. In addition, computational predictions did not capture the full complexity of splicing disruption observed for *POLR3A* or *OPA1*.

These observations highlight both the utility and the limitations of current *in silico* tools. While they are valuable for variant prioritisation, they are insufficient for definitive classification, particularly for variants located outside canonical splice sites or within 1–2 nucleotides of exon–intron boundaries, where predictive performance remains variable. In our cohort, functional evidence generated by Amp-LRS enabled application of the PVS1 criterion under ACMG/AMP guidelines, allowing reclassification of four variants from VUS to pathogenic or likely pathogenic (***Table 2***), a level of diagnostic certainty that could not have been achieved through *in silico* prediction alone.

Several limitations of our amplicon-based strategy should be acknowledged. First, this approach is restricted to genes expressed in accessible tissues such as blood or skin fibroblasts. We cannot exclude the possibility that additional or more pronounced splicing abnormalities occur in disease-relevant tissues, particularly the nervous system, or that tissue-specific isoforms are missed. This may be especially relevant for variants such as those in *POLR3A*, where splicing effects are known to be tissue-dependent [11]. Second, the use of primers targeting the 5′ and 3′ UTRs of the canonical transcript means that alternative transcription start sites or alternative transcript termini are not captured. Finally, as a PCR-based method, Amp-LRS is subject to amplification bias, potentially favouring shorter or more abundant transcripts [22], and precise isoform quantification using nanopore sequencing remains affected by a degree of technical noise [23].

Despite these limitations, functional validation of non-coding variants remains one of the major bottlenecks in the clinical interpretation of WGS data. Advances in long-read transcriptome analysis are beginning to address this gap. Our study highlights the value of targeted long-read sequencing of full-length transcripts as a cost-effective and versatile approach to elucidate the functional consequences of candidate splice-altering variants. This strategy is particularly well suited for variants that match the clinical phenotype but remain classified as VUS in the absence of functional evidence, and it has clear potential to improve diagnostic yield in rare neurological diseases.

## Data Availability

All data produced in the present study are available upon reasonable request to the authors

## Acknowledgments/Funding

The work is supported by Charcot-Marie-Tooth Association (SR-202504 to AC), AFM-Telethon (28813 to AC), European Research Council Starting Grant (101165557 to AC), National Ataxia Foundation, Medical Research Council (MR/T001712/1 to AC), Muscular Dystrophy UK (24GRO-PG24-0719-1 to JNS and AC). RC was supported by a Guarantors of Brain post-doctoral fellowship. AB thanks the European Academy of Neurology (EAN) for support via a Research fellowship 2025. GA thanks the Peripheral Nerve Society for support via a Clinical Training Fellowship 2025. SM thanks Ricerca Corrente RCR-2025 to Fondazione IRCCS Istituto Neurologico Carlo Besta for grant support. JNS is supported by Medical Research Council (MR/Y010949/1).

